# Stress Ulcer Prophylaxis Prescribing Options for Critically Ill Patients in Hospitals Participating in a Randomized Controlled Trial: A Protocol and Statistical Analysis Plan

**DOI:** 10.1101/2025.08.12.25333521

**Authors:** Akira Kuriyama, Nicole Zytaruk, Deborah Cook

## Abstract

**Background:** Proton pump inhibitors (PPIs) and histamine-2 receptor antagonists (H2RAs) predominate as stress ulcer prophylaxis (SUP) agents in intensive care units (ICUs). Prescribing patterns may change over time as randomized controlled trials evaluate SUP in critically ill patients.

**Objective:** This study aims to compare site-specific, self-reported SUP prescribing practices before and after the REVISE trial in participating ICUs.

**Methods:** This is a pre-planned study as part of the REVISE trial (NCT03374800), which compared intravenous pantoprazole with placebo in mechanically ventilated patients. The REVISE trial showed that intravenous pantoprazole reduced the risk of clinically important upper gastrointestinal bleeding (GIB) and patient-important upper GIB, without a significant effect on mortality or other clinical outcomes. We conducted a survey of all research teams (paired research coordinators and site physicians) in 68 participating trial centers, both before they enrolled patients and after the completion of the trial to record their self-reported SUP practice patterns.

**Results:** We will report ICU-level data on the typical SUP agent prescribed, the presence and content of preprinted orders or electronic admission order sets, and SUP discontinuation practices. We will apply descriptive statistics to summarize these patterns and compare them between the pre- and post-trial phases.

**Conclusions:** This study will describe contemporary data on SUP prescribing and discontinuation patterns before and after an international randomized controlled trial in participating trial centers.

## Background

Critically ill patients are at risk of developing upper gastrointestinal bleeding, with reported prevalence ranging from 1% to 9% (1). Although relatively uncommon, clinically important upper GIB may confer increased length of stay in the intensive care unit (ICU) or increase the risk of death (2). The predominant first-line stress ulcer prophylaxis (SUP) in ICUs has been proton pump inhibitors (PPI), followed by histamine-2 receptor antagonists (H2RA)(3, 4). In a study exploring temporal changes in SUP prescribing patterns in adult ICUs in the UK between 2020 and 2024, PPI prescriptions increased from 44.7% to 97.8%, whereas H2RA prescriptions declined from 49.4% to 0.4% (5)

Findings from large rigorous randomized controlled trials are expected to influence clinical practice. This may involve affirming current practice or changing current practice. Previous studies have shown mixed results regarding an expectation of practice change following randomized trials. Some trials with a neutral effect have paradoxically been followed by an observed increased use of the interventions studied (6) while the adoption of interventions shown to be beneficial in other trials appears to occur either rapidly (7-10) or gradually (11). Further, the deadoption of interventions in trials that have been shown to be harmful may be considerably delayed (12, 13).

A recent international randomized trial, <u>R</u>e-*Ev*aluating the <u>I</u>nhibition of <u>S</u>tress <u>E</u>rosions (REVISE), compared intravenous pantoprazole, a PPI, with placebo in 4,821 patients undergoing invasive mechanical ventilation (14). Pantoprazole was prescribed in the ICU until discontinuation of invasive mechanical ventilation, the occurrence of a prespecified indication or contraindication to PPI use, or death, up to 90 days. The trial showed that pantoprazole significantly reduced the risk of clinically important upper GIB and patient important upper GIB, without a significant effect on mortality or other clinical outcomes.

The overall objective of this study is to compare site-specific SUP self-reported prescribing before and after the REVISE Trial in participating ICUs. Specific aims are: (1) to examine prescribing options that include PPIs; and (2) to describe the patterns of SUP discontinuation.

## Methods

As part of our embedded knowledge translation work for the REVISE trial, we surveyed all research teams (paired research coordinators and site physicians) in 68 participating centers, both before they enrolled patients and after the completion of the trial (in May-June 2025), to record their self-reported SUP practice patterns. Research coordinators and physician investigators were selected as respondents due to their firsthand knowledge of SUP for patients enrolled and not enrolled in REVISE, before, during and after the trial conduct, due to daily screening, familiarity with local order sets and local discussions. In the pre- and post-trial surveys, the data included ICU-level information on the types of SUP prescribed, the presence or absence of pre-printed orders (PPOs) or electronic order sets, with associated drug class, and SUP discontinuation patterns. The predefined options were:

1. Type of SUP typically prescribed for most ICU patients (H2RA, PPI, sucralfate, antacid, no SUP, other (to specify))
2. Presence of preprinted orders or electronic admission order sets in the ICU (yes or no). If “Yes,” which SUP options were assessed (H2RA, PPI, sucralfate, SUP not included in admission order set, discontinuation when enteral feeds started, discontinuation when extubated, other (to specify)
3. Typical SUP discontinuation practices with multiple selections allowed (discontinuation when enteral feeds are fully tolerated, discontinuation when extubated, discontinuation upon ICU discharge, other (to specify)) Our hypotheses are as follows:
  1. **Use of PPI as the typical SUP agent** Hypothesis: The use of PPIs as the typical SUP may have increased after trial participation.
  2. **Discontinuation practices** Hypothesis: Discontinuing SUP at the time of extubation may have become more common after trial participation.
  3. **PPO drug classes** Hypothesis: More centers may have PPOs or electronic order sets, and the option for PPIs may have become more prominent therein after trial participation.

## Analysis

Descriptive statistics will be used to summarize practice patterns. We will report (mean (standard deviation) or median (interquartile range) as appropriate for continuous variables, and report frequency (and percentages) for categorical data.

We will use Fisher’s exact test or the Chi-square test, as appropriate, to compare binary outcomes between the two pre- and post-trial phases. We will use an unpaired Student’s t-test to compare continuous outcomes. We will define statistical significance at p<0.05.

## Ethics

Research ethics committees and relevant regulatory bodies approved the REVISE trial in all participating sites. Three informed consent models were used across 68 centers: *a priori* consent, deferred consent, and an opt-out model of enrolment in one center.

This pre-planned study to compare site-specific SUP self-reported prescribing before and after the REVISE Trial in participating ICUs is being submitted to the Hamilton Integrated Research Ethics Board (HiREB), which was the board of record for REVISE under Clinical Trials Ontario. No additional data are required from medical charts, research charts or research teams in any institution. No patients or families will be contacted. No patient-level data will be analyzed; only anonymized aggregate data will be presented at the institutional level.

## Discussion

The findings from this study will add to the literature by uniquely evaluating ICU-level SUP practice options and patterns before and after centers’ participation in the REVISE Trial.

Our study has several strengths. The center participation rate in this survey was 100% and there are no missing data for this analysis. Several surveys of SUP practice patterns have used questionnaires eliciting stated practice of individual clinicians (5, 15-22) (Table 1); others have employed observational or audit-based approaches to document actual practice (3, 10, 23-29) (Table 2). However, our study will exclusively focus on ICU-level prescribing options in centers that enrolled patients in a trial to help generate contemporary evidence about acid suppression.

**Table 1.**
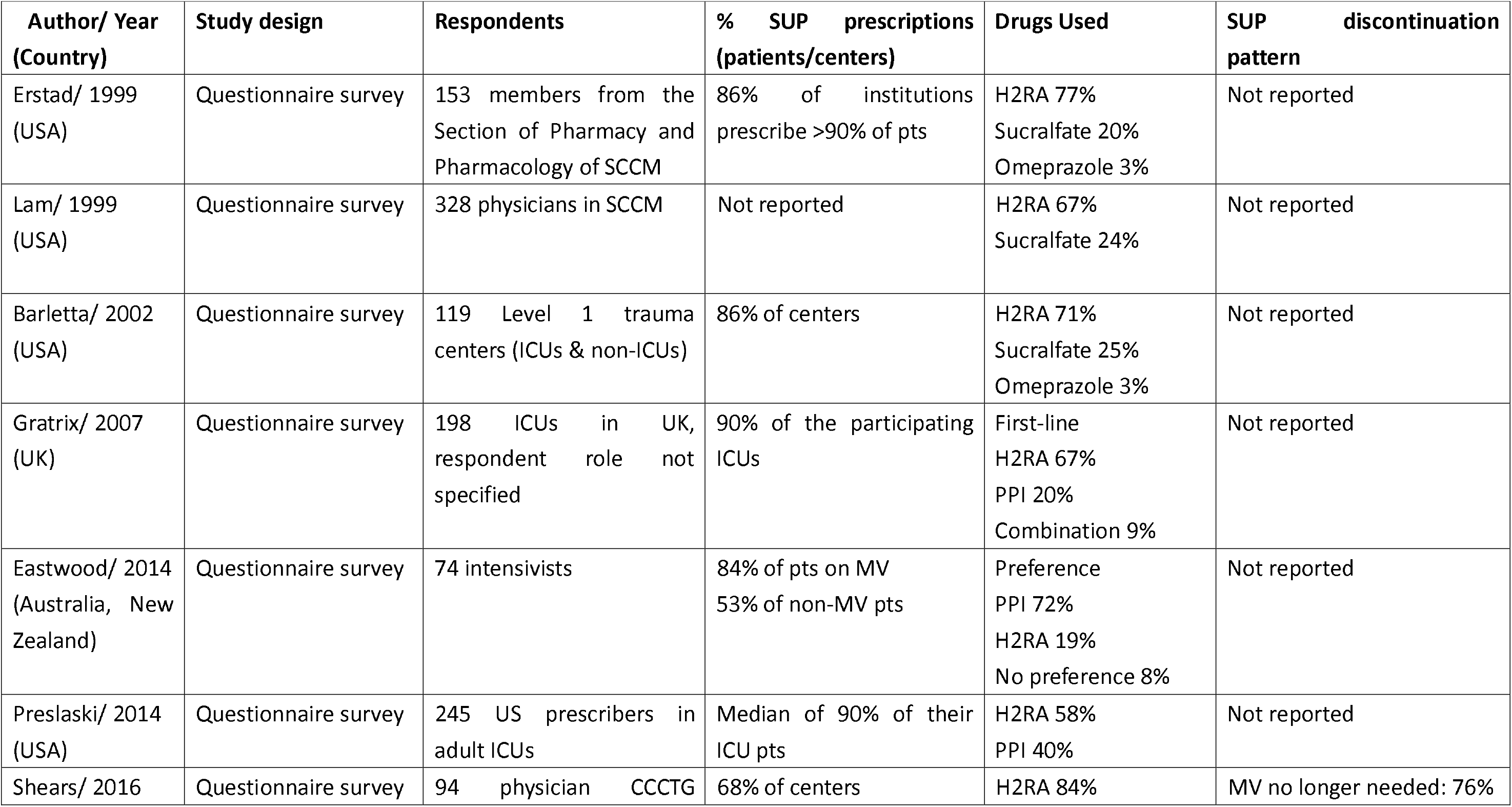
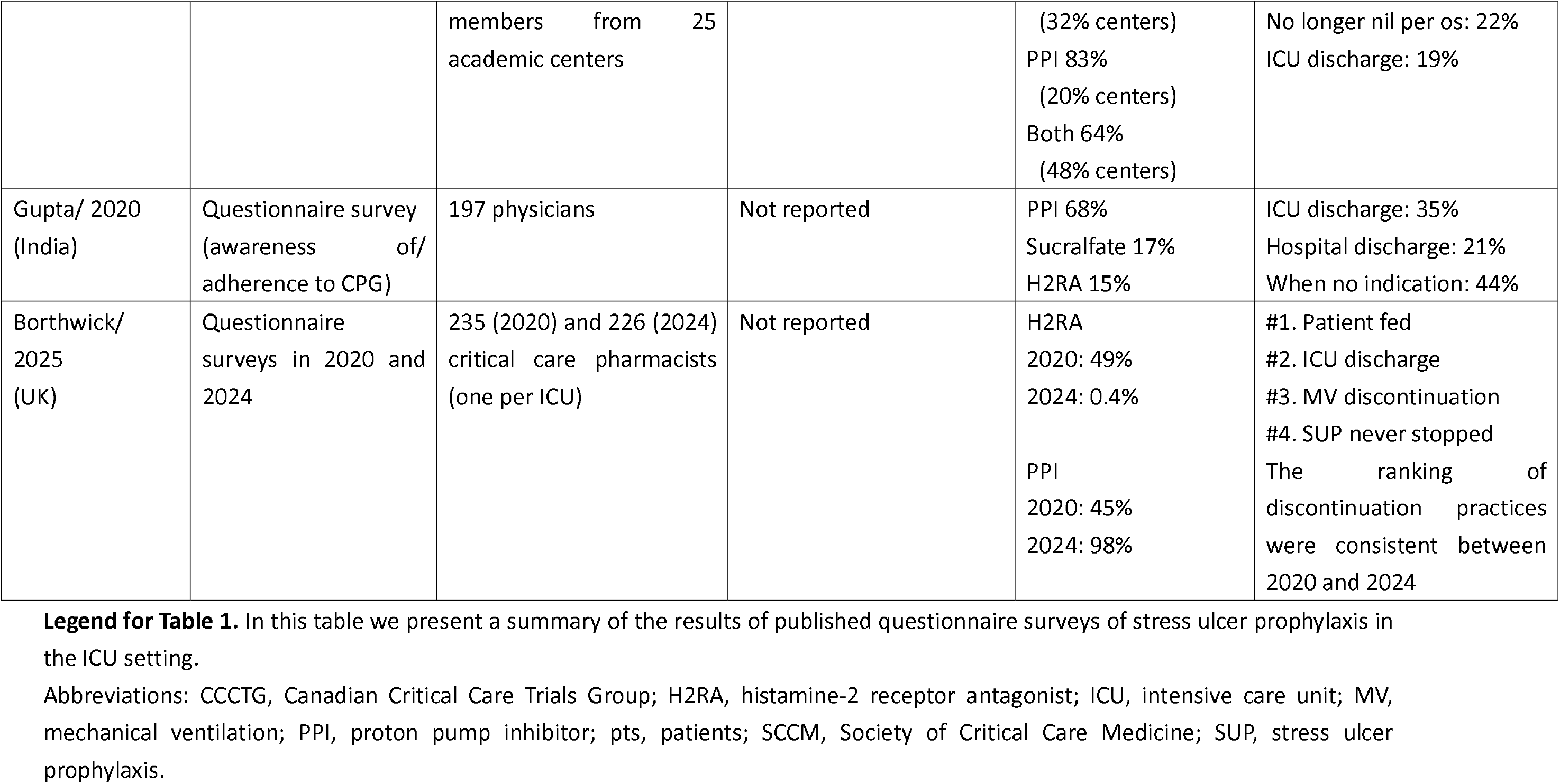
Summary of published questionnaire surveys of stress ulcer prophylaxis in the intensive care unit.

**Table 2.** Summary of published observational studies of stress ulcer prophylaxis in the intensive care unit.

| **Author/ Year**<br>**(Country)** | **Study design** | **Populations/ Settings** | **% pts on SUP** | **Drugs Used** | **SUP discontinuation**<br>**pattern** |
| --- | --- | --- | --- | --- | --- |
| Farley/ 2013<br>(Australia) | Retrospective chart<br>review | 387 adult pts in 2 ICUs | 85% of pts | PPI 95%<br>Ranitidine 5%<br>PPI & ranitidine<br>0.3% | Not reported |
| Barletta/ 2014<br>(Canada, USA) | Prospective<br>point-prevalence<br>study | 584 pts in 58 ICUs | 84% of pts | PPI 70%<br>H2RA 30% | Not reported |
| Franchitti/ 2020<br>(Switzerland) | Retrospective audit | 140 pts in an adult ICU | 93% of pts<br>(80% pt-days) | Esomeprazole 86%<br>Ranitidine 14%<br>Both 0.2%<br>(Calculated in<br>pt-days) | Not reported |
| Biyase/ 2021<br>(South Africa) | Retrospective, chart<br>review (adherence to<br>CPG) | 174 pts in 3 ICUs | 90% of pts | H2RA 51%<br>PPI 31%<br>Sucralfate 17%<br>H2RA & PPI 0.6% | Not reported |
| de Oliveira/<br>2021<br>(Brazil) | Retrospective<br>(adherence to CPG) | 105 pts in 3 general ICUs | 95% pt-days | Not reported | Not reported |
| Aramesh/ 2022<br>(Iran) | Prospective<br>observational study<br>(adherence to CPG) | 200 pts receiving SUP in a<br>general ICU | Not reported | H2RA 62%<br>PPI 38% | Not reported |
| McGraw/ 2024<br>(USA) | Retrospective study | 407 pts with TBI or SCI in<br>ICUs of 6 Level-1 trauma<br>centers | 83% of pts | H2RA 88%<br>PPI 14% | Not reported |
| Zhang/ 2024<br>(China) | Retrospective<br>(adherence to CPG) | 651 pts in an ICU | 59% of pts | PPI, majority (no<br>details reported) | Not reported |

**Legend for Table 2:** In this table we present a summary of published observational studies documenting stress ulcer prophylaxis patterns in the ICU setting.
Abbreviations: CPG, clinical practice guidelines; H2RA, histamine-2 receptor antagonist; ICU, intensive care unit; MV, mechanical ventilation; PPI, proton pump inhibitor; pts, patients; SCCM, Society of Critical Care Medicine; SCI, spinal cord injury; SUP, stress ulcer prophylaxis; TBI, traumatic brain injury.

Notwithstanding, our study has limitations. First, any associations or patterns observed including in relation to the REVISE trial will be interpreted cautiously as this study design precludes causal inferences. Our pre-post study is neither an audit of actual practice (3, 10, 23-29) nor survey of stated practice (5, 15-22) but will provide aggregate ICU-level prescribing options, without precisely capturing individualized approaches at the patient level, as recommended by practice guidelines (30). Third, having participated in a recent large trial, ICUs in this study may be more research-intensive than some others, limiting the generalizability of our findings. While 68 centers in 8 countries participated in this study, the results will not reflect universal practice.

## Data Availability

All data produced in the present work are contained in the manuscript.

